# COVID-19 Pandemic: Is Chronic Inflammation a Major Cause of Death?

**DOI:** 10.1101/2020.05.12.20099572

**Authors:** Albina Tskhay, Alena Yezhova, Kenneth Alibek

**Author notes:** Corresponding Author (Albina Tskhay).

## Abstract

**Background:** Today humanity is facing another infectious threat: a newly emerging virus SARS-CoV-2 causing COVID-19. It was already described that COVID-19 mortality among elderly people and people with such underlying conditions as obesity, cardiovascular diseases, cancer, chronic respiratory diseases, and diabetes s increased. Dysregulation of the immune responses vital for antiviral defense, which are typical for chronic inflammation, led us to a hypothesis that chronic inflammation is the main risk factor for increased susceptibility and mortality from COVID-19.

**Method:** Based on the available information for 126 countries, statistical analysis to find out whether the difference in incidence and mortality within countries can be explained by the existing chronic inflammation among the countries’ population, was conducted.

**Results:** A positive correlation between the percentage of people dying from chronic noncommunicable diseases and COVID-19 incidence (p<0.001) and mortality (p<0.001) within countries.

**Conclusion:** The problem of COVID-19-caused high mortality rate may be a consequence of the high number of people having chronic low-grade inflammation as a precondition, and thus, one of the potential ways to reduce risk of morbidity and mortality is to focus on this widespread health problem, mainly occurring in developed countries and to take corresponding diagnostic, preventative, and treatment measures.

## Introduction

Currently, we are facing a new infectious threat: recently emerged virus SARS-CoV-2 causing coronavirus disease (COVID-19), a disease that has already killed about 270,000 people worldwide in just four months. SARS-CoV-2, a β-CoV, is the seventh coronavirus that has become pathogenic to humans through the process of zoonosis, causing COVID-19, a potentially fatal disease that is of great concern to the global public health and the economic development of every country in the world (Rothan and Byrareddy, 2020). The symptoms of COVID-19 infection appear after an incubation period of approximately 3 to 5 days and include fever, cough, fatigue, sputum production, headache, hemoptysis, etc. (Guan et al. 2020).

It can be noticed that despite similar countermeasures taken in almost all countries, which include self-isolation, quarantine, social distancing, and wearing face masks, the rates of COVID-19 incidence and mortality differ significantly among countries. There are numerous hypotheses about the underlying causes for the observed differences, but as the research on this continues, it is becoming obvious that the main factor is the immune system function and response to the virus among countries’ populations.

Specific observation described was increased COVID-19 mortality among elderly people and people with underlying conditions such as obesity, cardiovascular diseases, cancer, chronic respiratory diseases, and diabetes (Novel Coronavirus Pneumonia Emergency Response Epidemiology Team 2020; Lighter et al. 2020). The median age of hospital patients who tested positive for COVID-19 is approximately 63 years (Rothan and Byrareddy, 2020). This situation is not unique, as it is a well-known fact that aging and the abovementioned diseases and conditions are accompanied by chronic inflammation and insufficiency and dysregulation of immune system response (Chung et al. 2019; Zhong and Shi 2019). This results in the immune system’s inability to fight infections (Boe et al. 2016; Oh et al. 2019). It is known that elevated levels of proinflammatory mediators (C-reactive protein, tumor necrosis factor, interleukins 1-beta and 6) are associated with infectious disease-caused mortality in these people (Boe et al. 2016). Apart from this, the elevation of proinflammatory mediators also negatively affects hemoglobin concentrations, insulin-like growth factor 1 levels, and levels of albumin, micronutrients, and vitamins (Chen et al. 2019).

In several works (Meyer 2010; Boe et al; 2016; Oh et al. 2019), the changes in the immune system observed in people with chronic low-grade inflammation were well summarized. Briefly, some of these changes include:

- Reduced phagocytic activity and number of monocytes and macrophages
- Reduction in MHC II expression
- Altered cytokines production
- Altered TLR expression and signaling
- Decreased natural killer (NK) cells cytotoxic activity
- Dysfunctional neutrophils
- Decreased number of naïve T and B cells
- Increased accumulation of senescent T cells
- Diminished response of T cells to new antigens

The combination of these immune abnormalities is named immuno-aging. This condition is characterized by dysregulation of both the innate and adaptive immune system, and the malfunctioning of the immune system in the respiratory system of these individuals is especially well described. Specifically, elevated levels of proinflammatory mediators contribute to diminished pulmonary function and dysregulate immune responses to respiratory infections which are characterized by reduced mucociliary clearance, upregulation of proteins participating in the attachment of pathogens to epithelial cells of the respiratory system, reduction of toll-like receptors (TLR) expression on lung cells, etc. (Boe et al. 2016). In addition to the numerous immune system alterations increasing the susceptibility to respiratory infections, existing lung inflammatory responses upon new infection additionally increase susceptibility to bacterial co-infections such as *Streptococcus pneumoniae* and others (Aguilera and Lenz 2020).

As a matter of fact, the initial immune response plays a crucial role in the COVID-19 outcome. Cells generally respond to virus infection by mounting an innate antiviral response to limit the spread of the infection and aid in inducing an adaptive immune response that will eventually clear the virus. For instance, coronaviruses exploit the toll-like and some other receptors for initial attachment. Activation of one or more of these sensors generally leads to the activation of the transcription factors interferon (IFN)-regulatory factors 3 and 7 and NF-κB. These stimulate the expression and excretion of type-I IFN and pro-inflammatory cytokines, which in turn activate the JAK-STAT signaling cascade that induces the expression of antiviral interferon-stimulated genes (ISGs). This ultimately results in an antiviral state of the infected and neighboring cells. ISGs were shown to target virtually all steps of the viral cycle in order to restrict viral replication (de Wilde et al., 2018). However, as it was found, the TLRs are downregulated in the condition of immunoaging (Oh et al. 2019). At the same time, the severity of coronaviral infection depends on the IFN response timing relative to infection. Immediate production of one’s own IFN-I or exogenous IFN administration can result in clearance of the virus or a delayed and slowed multiplication, leading either to the termination of infection by the innate mechanisms or to the elimination of the virus by timely or already formed mechanisms of specific immunity. On the other hand, a delay in IFN-I response will most likely result in the increase of pro-inflammatory cytokine production and more severe outcomes (Channappanavar et al. 2019).

Dysregulated immune responses could potentially drive the COVID-19 hallmark syndromes such as acute respiratory distress syndrome (ARDS), cytokine release syndrome (CRS), and lymphopenia. The initial mode of viral pathogen-associated signal (PAMP) recognition by innate cells has a major impact on downstream myeloid signaling and cytokine secretion. Significantly elevated systemic levels of pro-inflammatory cytokine IL-6 have been reported in several COVID-19 patient cohorts and shown to correlate with disease severity. Increased IL-6 can also be associated with higher levels of IL-2, IL-7, IFN-α, and GM-CSF as seen in secondary hemophagocytic lymphohistiocytosis. In response to viral infections, mononuclear phagocytes (MNPs) drive interleukin, and IFN-I and IFN-III production, resulting in inflammasome activation, induction of pathogenic Th1 and Th17 cell responses, recruitment of effector immune cells and cytokine release syndrome pathology (Vabret et al., 2020).

Dysregulation of the immune responses vital for antiviral defense, which are typical for chronic inflammation, led us to the hypothesis that chronic inflammation is the main risk factor for increased COVID-19 morbidity and mortality. Based on available information, in this work, we aimed to conduct a statistical analysis to find out whether the difference in incidence and mortality within countries can be predominantly explained by the chronic inflammation among the countries’ population.

## Methods

To assess the chronic inflammation incidence among populations, the data from the World Health Organization available from https://www.who.int/nmh/countries/en/ was used. In these country profiles collected in 2018, the data on the percentage of people dying of chronic diseases are reported. Countries for which these data were not reported were excluded from the report. Data of COVID-19 cases and death per million of population for each country were obtained from https://www.worldometers.info/coronavirus/ in the afternoon (EST) of May 2nd, 2020. Countries with the data absent were excluded from the analyses. The information about the incidence of COVID-19 and mortality from these diseases as well as the information about the death rate due to chronic noncommunicable diseases was available for 126 countries.

Pearson’s and Spearman’s correlation statistical analyses were conducted using JASP (2020). For more reliable results, additionally, Vovk-Sellke Maximum p-ratio (VS-MPR) also was included in the analyses (Sellke et al. 2011).

## Results

The statistically significant correlation between the percentage of people dying of chronic diseases and the number of COVID-19 caused deaths per million of population in each country as a result of using both Pearson’s and Spearman’s correlations methods with P-value <0.001 and very high VS-MRPs (Tables 1 and 2). It was noted that the COVID-19 mortality rate is not increasing gradually with the chronic diseases mortality but rather there is a sharp increase observed in the countries where chronic diseases mortality is higher than 80% (Figure 1a)

**Table 1.**
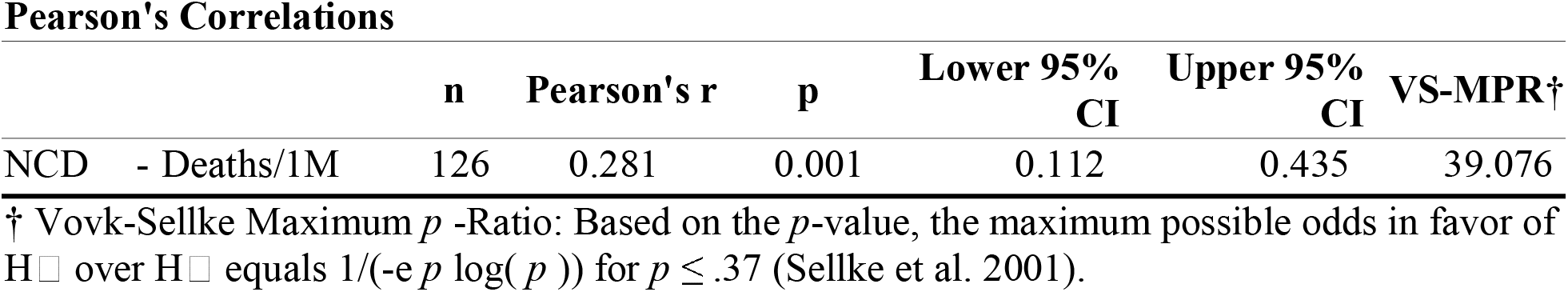
Pearson’s correlations between noncommunicable chronic diseases death rate and COVID-19 mortality among countries.

**Table 2.** Pearson’s and Spearman’s correlations between noncommunicable chronic diseases death rate and COVID-19 mortality among countries.

|  | n | Spearman's rho | p | Lower 95% CI | Upper 95% CI | VS-MPR |
| --- | --- | --- | --- | --- | --- | --- |
| NCD - Deaths/1M | 126 | 0.628 | < .001 | 0.509 | 0.724 | 3.210e<sup>+</sup>12 |

**Figure 1.**
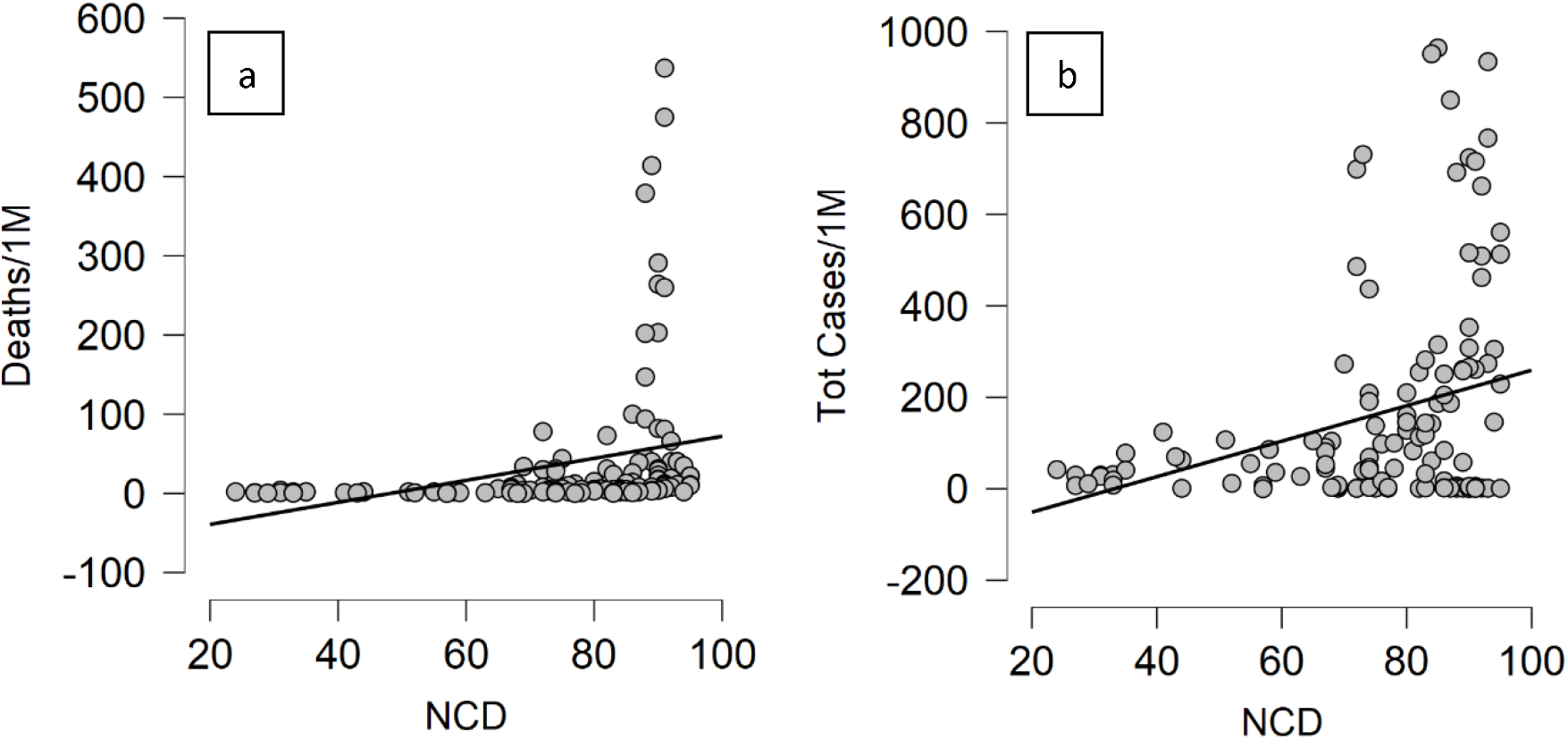
Graphical representation of correlations between noncommunicable chronic diseases death rate and COVID-19 mortality (a) and incidence (b) among countries.

The statistically significant correlation between the percentage of people dying of chronic diseases and the incidence of COVID-19 per million of population in each country was a result of using only Pearson’s correlations method with a p-value <0.001 and very high VS-MRP as well (Table 3). In Figure 1b, it can be seen that the incidence is gradually increasing with the percentage of people having chronic diseases with considerable difference observed when the percentage of people dying of chronic diseases is 70% and higher (mostly high-income countries).

**Table 3.** Pearson’s correlations between noncommunicable chronic diseases death rate and COVID-19 incidence among countries.

|  | n | Pearson's r | p | Lower 95% CI | Upper 95% CI | VS-MPR† |
| --- | --- | --- | --- | --- | --- | --- |
| NCD - Tot Cases/1M | 126 | 0.304 | < .001 | 0.136 | 0.454 | 89.189 |

## Discussion

Chronic noncommunicable diseases are a known risk factor for susceptibility to COVID-19 and mortality from this disease. Additionally, statistical analysis showed that overall patterns of incidence and mortality in countries also may depend on the chronic diseases’ prevalence. This could mean that most COVID-19 victims even if they are not affected by cancer, cardiovascular diseases, or other comorbidities associated with increased COVID-19 mortality, are already affected by chronic low-grade inflammation and resulting from this, immunoaging. It is not widespread knowledge, but a large proportion of population are carriers of chronic respiratory bacterial infections such as *Neisseria meningitidis, Haemophilus influenzae*, and *Staphylococcus aureus*. (Bhatta et al. 2014; Peterson et al. 2014). For instance, in some countries, the overall asymptomatic chronic carriage of *N. meningitidis*, a respiratory pathogen, reaches 60% to 70% (Bidmos et al. 2011; Rizek et al. 2016). This is especially important to pay attention to because it was reported that COVID-19 patients with higher neutrophil-to-lymphocyte ratio usually tend to have higher severity and mortality rate (Liu et al. 2020). While increased neutrophils and decreased lymphocytes are more typical for bacterial infection, this indicates possible bacterial-viral co-infection which increases the risk of a fatal outcome for patients with a pre-existing chronic bacterial infection.

Chronic inflammation is also likely to create another challenge for fighting COVID-19.

Currently, the main attempts on finding a solution are focused on vaccine development while it is known that chronic inflammation and resulting immune system dysregulation reduce the immunogenicity of vaccines. As it was already shown in animals in the case of SARS-CoV, similar coronavirus infection leading to atypical pneumonia, young mice immunized with anti-SARS vaccine appeared to be protected, but aged mice (the ones with immunoaging) infected with SARS demonstrated more severe symptoms than unvaccinated mice (Deming et al. 2006).

The problem of chronic diseases is not just a factor affecting survival from COVID-19, but also overall longevity of populations since chronic noncommunicable diseases are the leading death cause in most countries. “Currently, chronic diseases are the leading cause of death in the world and their impact is steadily growing. Approximately 17 million people die prematurely each year as a result of the global epidemic of chronic disease” (WHO 2019).

This indicates a critical need for searching for solutions to reduce chronic inflammation. Although chronic diseases we discuss here are also called chronic noncommunicable diseases, there is still an accumulated knowledge on the role of chronic persistent and latent bacterial and viral infections in these diseases (Knobler 2004; O’Connor et al. 2006; Ogoina and Onyemelukwe 2009).

Numerous studies, case reports, and clinical trials have already shown that the treatment of infections may have a highly beneficial health effect in relation to noncommunicable diseases. For instance, antiviral therapy inhibited tumors in cases of lymphoma and leukemia (Abdulkarim and Bourhis 2001; Hermine et al. 1995). MALT lymphoma and gastric cancer cells proliferation, in many cases caused by H. pylori, can be eliminated or reduced in most cases when treated with antibacterial therapy (Andriani et al. 2009; Tang et al. 2014). Antiviral drugs were shown to reduce cell proliferation in breast cancer (Pettersson et al. 2011). Treatment of glioblastoma patients with an antiviral drug was shown to reduce tumor growth by 72%, increase the number of patients with 2-year survival by 72% and increase life expectancy by 42 months (13.5 vs 56 months) (Söderberg-Nauclér et al. 2013). 80% of hepatocellular carcinomas are associated with HBV and HCV, and every year 500,000 lives can be saved if these viruses are treated using antiviral therapy (Alibek et al. 2012). Antiviral drugs were shown to decrease tumor size and to induce apoptosis in HPV-caused nasopharyngeal cancer (Yoshizaki et al. 2008). In patients with diabetes type 2, the use of antibiotics improved glycemic control, decreased the levels of inflammatory markers, and reduced the severity of insulin resistance (Bharti et al. 2013). As well, it was shown that antibacterial treatment soon after the infection reduced the risk of atherosclerosis development by 85% (Fong 2000). However, despite these findings, none of these noncommunicable diseases are treated using antimicrobial therapies.

Realizing that we cannot guarantee that our suggestions can help in solving all problems, our efforts can be focused on three directions with some or high probability of reducing risks:

1. Clinics in many countries do not include main inflammatory markers (such as C-reactive protein) to the routine tests (Bazeley et al. 2011). The widespread testing of known markers and the creation of new diagnostic approaches and methods to assess systemic inflammation or its individual branches will help to prevent the emergence of chronic disease or allow it to be treated in the early stages. Even though there is no universally established criterium, these tests below can help in detecting the pro-inflammatory state.

- Blood and/or saliva

- C-reactive protein
- IL-6
- Ferritin level
- TNF-alpha
- Fibrinogen level
- Blood:

- Hemoglobin A1C
- Erythrocyte count and sedimentation rate
- Lymphocyte count
- Neutrophil count
- White blood cell count
- Creatine kinase
2. There is a need to create a new understanding and principles of treatment with existing and new antibacterial and antiviral drugs, as the concept of short-term use of both should be revised. Chronic infections cannot be cured by short treatment courses. Moreover, in view of the so-called poly-infection or burden of infections, it is necessary to begin to consider the possibility of combined therapies, such as antibacterial and antiviral drugs administered simultaneously.
3. Despite the already relatively well-established understanding of chronic inflammation, medical science, and practice for the treatment of chronic inflammation still do not exist. The list of drugs for the treatment of these pathologies is very narrow, and the creation of integrated approaches to anti-inflammatory therapy combined with effective anti-infectious treatment can significantly prevent premature deaths promoted by the abovementioned conditions.

## Conclusion

Although life expectancy in high-income countries is significantly higher than in the developing ones, there is still a health problem to be targeted to increase longevity and improve life quality – chronic inflammation. The COVID-19 pandemic revealed an underestimation of chronic inflammation playing role in increased mortality in the majority of developed countries, and this problem is very likely to be detrimental for survival during the COVID-19 pandemic. In order to reduce the risk of death resulting from COVID-19 and other unexpected health threats, there is a necessity to reconsider more comprehensive methods of chronic inflammation diagnosis and prevention and treatment of these conditions.

## Data Availability

Not applicable

## Acknowledgments

We are grateful to Andrew Lefkowitz, CEO and chairman of FLAASK, LLC, for his financial, administrative, and moral support provided for this work.

## Declarations

Ethics approval and consent to participate – not applicable

Consent for publications – not applicable

Competing interests - authors have no competing financial, professional, or personal interests that might have influenced the performance or presentation of the work described in this manuscript.

